# Bayesian Calibration of Using CO_2_ Sensors to Assess Ventilation Conditions and Associated COVID-19 Airborne Aerosol Transmission Risk in Schools

**DOI:** 10.1101/2021.01.29.21250791

**Authors:** Danlin Hou, Ali Katal, Liangzhu (Leon) Wang

## Abstract

Ventilation rate plays a significant role in preventing the airborne transmission of diseases in indoor spaces. Classrooms are a considerable challenge during the COVID-19 pandemic because of large occupancy density and mainly poor ventilation conditions. The indoor CO_2_ level may be used as an index for estimating the ventilation rate and airborne infection risk. In this work, we analyzed a one-day measurement of CO_2_ levels in three schools to estimate the ventilation rate and airborne infection risk. Sensitivity analysis and Bayesian calibration methods were applied to identify uncertainties and calibrate key parameters. The outdoor ventilation rate with a 95% confidence was 1.96 ± 0.31ACH for Room 1 with mechanical ventilation and fully open window, 0.40 ± 0.08 ACH for Rooms 2, and 0.79 ± 0.06 ACH for Room 3 with only windows open. A time-averaged CO_2_ level < 450 ppm is equivalent to a ventilation rate > 10 ACH in all three rooms. We also defined the probability of the COVID-19 airborne infection risk associated with ventilation uncertainties. The outdoor ventilation threshold to prevent classroom COVID-19 aerosol spreading is between 3 – 8 ACH, and the CO_2_ threshold is around 500 ppm of a school day (< 8 hr) for the three schools.

**Practical Implications:** The actual outdoor ventilation rate in a room cannot be easily measured, but it can be calculated by measuring the transient indoor CO_2_ level. Uncertainty in input parameters can result in uncertainty in the calculated ventilation rate. Our three classrooms study shows that the estimated ventilation rate considering various input parameters’ uncertainties is between ± 8-20 %. As a result, the uncertainty of the ventilation rate contributes to the estimated COVID-19 airborne aerosol infection risk’s uncertainty up to ± 10 %. Other studies can apply the proposed Bayesian and MCMC method to estimating building ventilation rates and airborne aerosol infection risks based on actual measurement data such as CO_2_ levels with uncertainties and sensitivity of input parameters identified. The outdoor ventilation rate and CO_2_ threshold values as functions of exposure times could be used as the baseline models to develop correlations to be implemented by cheap/portable sensors to be applied in similar situations to monitor ventilation conditions and airborne risk levels.

## 1. Introduction

Airborne transmission of relatively small aerosol droplets plays a dominant role in spreading SARS-CoV-2 (hereafter as COVID-19), especially in indoor spaces ^1,2^. School classrooms pose a considerable challenge because of the large occupants’ density, mandatory presence of students, and ventilation conditions of concern ^3^. For example, in Quebec province, Canada, a partial lockdown was in effect for non-essential business, with many offices closed whereas primary and secondary schools open. The weekly school-related COVID-19 cases in Quebec at the end of August 2020 showed that schools accounted for 20% of the province’s COVID-19 cases, while students and staffs account for about 18% of Quebec’s population ^4^. Statistical data shows that 1,781 schools had been observed with at least one positive case in Quebec since the beginning of the pandemic^5^. Therefore, the rate of COVID-19 transmission in schools was higher than the community transmission, and mitigation measures must be implemented in classrooms to reduce the infection risk. Several studies revealed the significant impact of ventilation rate in reducing or preventing the airborne transmission of diseases in indoor environments ^6^. There are different recommendations for the minimum required ventilation rate in indoor spaces to achieve an acceptable indoor air quality or preventing indoor airborne transmission. The United States Centers for Disease Control and Prevention (CDC) and World Health Organization (WHO) recommended a minimum ventilation rate of 12 ACH to prevent airborne transmission in health-care facilities ^7,8^. The Harvard-CU Boulder Portable Air Cleaner Calculator ^9^ suggests a total of five air changes per hour as a good ventilation condition for reducing airborne transmission risk in classrooms.

While recommendations are mainly based on the ventilation rate, it has been a challenge to quantify the outdoor air ventilation rate in a room. Indoor air CO_2_ concentration is often considered a surrogate/indicator for the ventilation rate. For example, the Montreal school board (Centre de services scolaire de Montreal) stated in an open letter on December 14, 2020: “Establishments without a mechanical ventilation system should apply the window opening guidelines to ensure frequent air changes in our premises”; “Always in order to ensure good indoor air quality, we have also started measuring carbon dioxide (CO_2_) in our establishments since November. In addition to this initiative, there are the CO_2_ tests that must be carried out by all school service centers in Quebec before December 16. The level of CO_2_ is a good indicator of the supply of fresh air in a room. Thus, following these tests, corrective measures will be put forward, if necessary.” ^13^.

In the literature, several studies used a transient CO_2_ mass balance method and measured CO_2_ levels to calculate the ventilation rate in different indoor environments such as classrooms and university libraries ^10–12^. Batterman (2017) ^12^ estimated the CO_2_ generation rate based on the age and assumed activity level for CO_2_ calculation in mechanically ventilated classrooms, but the real activity type was unknown, so the results were subject to uncertainties. They used the whole day data to estimate ventilation rate, but they did not validate the model by calculating the CO_2_ at a different time or day. For the estimation of ventilation rate using the transient CO_2_ sensor data, it is essential to find dominant parameters that affect the final results, calibrate the model by measurement data, quantify and report the ventilation rates and infection risks with uncertainties. As a result, due to various factors affecting CO_2_ levels, such as variable occupant numbers and outdoor conditions, and the fact the unknown uncertainties of these factors, questions have been raised about using CO_2_ sensors to assess COVID-19 transmission risk ^14^. In summary, therefore, two questions are often raised: 1). The CO_2_ for Ventilation Assessment Question: when considering variable occupancy and dynamic surrounding environment, how can we relate CO_2_ concentrations to ventilation rates with uncertainties taken into account? 2). The CO_2_ for Risk Assessment Question: If CO2 is used as an indicator for the COVID-19 airborne aerosol transmission risk associated with ventilation conditions, what are the CO_2_ threshold levels to prevent spreading for ventilation-related risks with uncertainties quantified?

Both questions center around the uncertainties and associated sensitivity analysis of parameters. Sensitivity Analysis (SA) and calibration methods such as Bayesian Markov Chain Monte Carlo (MCMC) method ^15^ can be used along with measured indoor CO_2_ concentrations to find the dominant parameters for the CO2 levels such as ventilation rate and calibrate them. Bayesian MCMC is a calibration technique proposed in the twentieth century owing to the development of MCMC and modern computer. Its application to the computer models’ calibration was systematically illustrated by Kennedy and O’Hagan (2001) ^16^. From then on, the boom of Bayesian inference and calibration was signified. Now, Bayesian inference calibration has been utilized in various topics, such as environment ^17–20^, hydrology ^21–23^, transportation ^24^, and medicine research ^25^. In the field of heating, ventilation, and air conditioning (HVAC), one of the early applications was conducted by Heo et al. ^26^. Building energy models were calibrated using Bayesian inference and further analyzed considering the uncertainties for retrofit decision making. By propagating parameters using probabilistic analysis, Bayesian inference incorporates uncertainties into real systems’ approximations by computer models. Combining multiple sources of information at different scales and with different reliabilities, the inadequacy of a model, revealed by the discrepancy between the predictions and observed data, can be corrected. Compared with conventional deterministic calibration methods, the advantages of Bayesian inference calibration are: 1) When the calibration measurements are qualitatively/quantitatively insufficient, for traditional methods, the estimated model parameters can be far off from their original values; however, for Bayesian calibration, since the uncertainties are considered, the calibration results are more stable and reasonable; 2) For the traditional calibration method, the results are often deterministic. In comparison, for the Bayesian inference calibration method, the results are derived from quantitative stochastic analysis and with possibilities that can be regarded as a degree of belief.

Therefore, to answer the “CO_2_ for Ventilation Assessment” question, in this work, we investigated three classrooms from three schools in Montreal, Canada, to analyze CO_2_ and ventilation rate, and estimate COVID-19 airborne aerosol infection risk. To estimate ventilation rate using the transient CO_2_ mass balance model, we obtained the measurement data of transient CO_2_ concentrations. We conducted a sensitivity analysis to find the dominant parameters for indoor air CO_2_ concentration. The Bayesian MCMC method was then used with the three classrooms’ measured CO_2_ data to calibrate the dominant parameters and quantify the uncertainties. Calibrated ventilation rate helps teachers to compare it with the officially recommended value for preventing indoor airborne transmission. Then we used the calibrated model and tried different ventilation rates to estimate CO_2_ level for different exposure times. The results help occupants to estimate the ventilation rate from the measured CO_2_ data at each time.

On the other hand, calculation and adjusting the ventilation rate may not be enough for mitigating the airborne transmission of COVID-19 in classrooms because no definite ventilation and CO_2_ thresholds have been agreed upon for COVID-19. Recommended ventilation rate or indoor air CO_2_ level by some standards and agencies for indoor air quality condition may not be enough to prevent the indoor airborne transmission. For example, ASHRAE Standard 62-2001 (ASHRAE, 2004) recommended a maximum CO_2_ level of 1000 ppm in classrooms for acceptable indoor air quality, which may still not be enough for preventing airborne transmission of diseases. The National Institute for Occupational Safety and Health (NIOSH) recommended a CO_2_ level of 600-1500 ppm for schools and workplaces but only considered comfort and working efficiency ^27^. An unofficial study by some scientists and teachers in 25 classrooms in Montreal revealed that in 75% of the classrooms, the level of CO_2_ exceeded 800 ppm because of poor ventilation conditions ^28^. However, they have been confused about the relations between monitored CO_2_ levels, ventilation conditions, and COVID-19 airborne transmission risks in the classrooms. The Wells-Riley model ^29^ can be used to calculate the indoor infection risk using calibrated ventilation rate. The effective reproductive ratio, R_0_ (ratio between secondary infectious cases and source cases), is often applied and should be less than one to prevent spreading. Du et al. (2020) studied the impact of ventilation improvement on a real tuberculosis (TB) outbreak in underventilated university buildings. Their result showed that improving indoor ventilation to levels correspond with CO_2_ < 1000 ppm reduced 97% of TB infection risk. Before improving the ventilation, the initial CO_2_ concentration was 3,204 ppm in one classroom where the index cases attended. They concluded that enhancing the ventilation rate to 14-15 ACH could effectively control the TB outbreak. However, there is no information given for the COVID-19 regarding the CO_2_ concentration for monitoring to prevent airborne aerosol spread.

Rudnick and Milton (2003) derived an equation to estimate the indoor airborne infection transmission using indoor air CO_2_ concentration. They calculated the critical rebreathed fraction of indoor air below which airborne propagation of typical respiratory infections will not occur. They modeled several hypothetical cases in their work without measurement data. Peng and Jimenez (2020) ^31^ derived an analytical expression of CO_2_-based risk proxies for COVID-19 and used it to estimate CO_2_ level corresponding to an acceptable airborne risk level in different indoor environments. They showed that acceptable CO_2_ level varies by over two orders of magnitude for various rooms and activities. It also depends on other factors, such as wearing face masks. No measurements and uncertainties were reported. Eykelbosh (2021) ^14^ reviewed several studies that used indoor CO_2_ level in assessing the transmission risk and concluded that indoor air CO_2_ level could only represent the ventilation condition. The infection risk does not depend only on the ventilation rate, and other factors such as wearing a face mask, using portable air cleaner, and exposure time can also affect the infection risk. Therefore, to estimate the required ventilation rate and critical CO_2_ level to prevent the transmission of COVID-19 aerosols in a classroom, it is crucial to know the actual room condition such as occupancy profile, activity type, and other parameters that affect the estimation of infection risk.

Therefore, to answer the “CO_2_ for Risk Assessment” question, in this work, we used the calibrated ventilation rate and actual room parameters to calculate the COVID-19 airborne transmission in the three classrooms using the modified Wells-Riley equation. We compared the results with infection risk corresponding to the Reproductive number to be one (R_0_ = 1), and different ventilation rates, and CO_2_ threshold levels at various exposure durations.

## 2. Methodology

This section presents the models of CO_2_ concentration, airborne infection risk, and sensitivity analysis, and finally, the Bayesian calibration methods. Two fully-mixed transient mass balance equations are solved to calculate indoor air CO_2_ and COVID-19 quanta concentration. The schematic of the models is plotted in Figure 1.

**Fig. 1.**
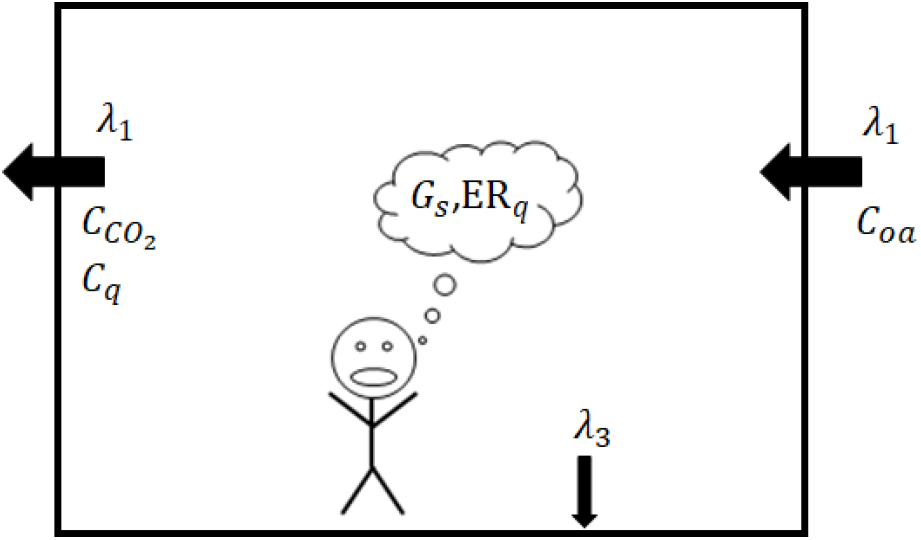
Schematic of the indoor air CO_2_ and quanta concentration models

### 2.1 Indoor CO_2_ concentration model

A fully-mixed transient mass balance model is solved for the calculation of CO_2_ concentration in the room.

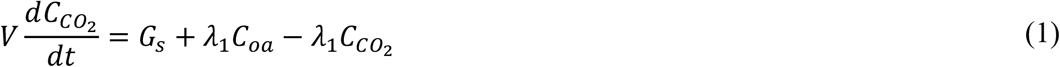

where *V* is the room volume 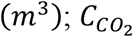 is the indoor air CO_2_ concentration (*mg*/*m*^3^); t is the time duration (*s*); *G*_*s*_ is the CO_2_ generation rate by all occupants (*mg*/*s*), which depends on the age and activity level; *λ*_1_ is the total outdoor air ventilation rate (*m*^3^/*s*); and *C*_*oa*_ is the outdoor air CO_2_ concentration (*mg*/*m*^3^).

The transient mass balance, Eq. 1, is useful for solving arbitrary occupancy patterns and generation rates such as classrooms because students leave the classroom for break and launch. The solution of Eq. 1 is:

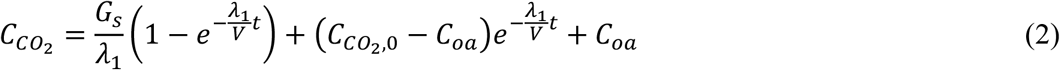

where 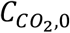 is the observed initial CO_2_ concentration at each occupancy phase.

### 2.2 Airborne aerosol infection risk model

The probability of infection (PI) of a susceptible person in the room is calculated using the Wells-Riley formulation^32^. The method was first used by Jimenez et al. ^33^ for calculating infection risk in different indoor environments and is recently applied to the City Reduced Probability of Infection (CityRPI) model and used for city-scale infection risk analysis ^34,35^. PI is a function of the number of quanta *μ* inhaled by the susceptible person (Eq. 3). We assumed that social distancing is maintained between all occupants, and the current study focuses on airborne aerosol transmission only. We used five assumptions for applying this model: i) there is only one infected person in the room who emits SARS-CoV-2 quanta with a constant rate, ii) the initial quanta concentration is zero, iii) the latent period of the disease is longer than the duration students stay in the classroom. Therefore the quanta emission rate remains constant during the day, iv) the indoor environment is well-mixed, and v) the infectious quanta is removed as a first-order process by the ventilation, filtration, deposition on surfaces, and airborne inactivation. The PI in Eq. 1 is based on the attendance of one infected person in the room, so it calculates the probability that COVID-19 aerosols are transmitted from the infected person to a susceptible person in the room; therefore, it is a conditional probability of infection (PI_cond_).

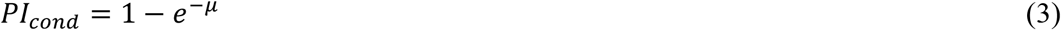

The number of quanta inhaled by the susceptible person at the exposure time T is calculated by time-averaged quanta concentration.

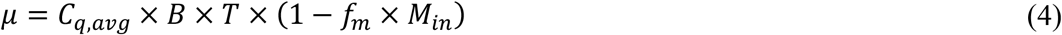

B is the inhalation rate (*m*^3^/*h*); *C*_*q,avg*_ is the time-average quanta concentration (*q*/*m*^3^); *T* is the exposure time (*h*); *f*_*m*_ is the fraction of people in the room who wears the mask, and *M*_*in*_ is the inhalation mask efficiency. A well-mixed transient mass balance equation similar to Eq. 1 is solved to calculate the room’s transient quanta concentration.

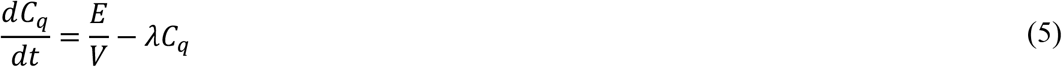

where *C*_*q*_ is the indoor quanta concentration (*q*/*m*^3^); *E* is the net quanta emission rate (*h*^−1^); and *λ* is the first-order loss rate coefficient for quanta (*h*^−1^). Assuming that the initial quanta concentration is zero at the beginning of the day, Eq. 5 is solved as follows:

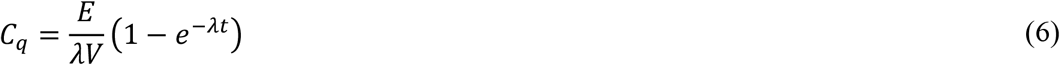

Because of the change in the occupancy pattern during the day, the time-averaged quanta concentration is calculated using the Trapezoidal integration. *E* is calculated based on the number of infected people in the room *N*_*inf*_, the fraction of people in the room with the mask *f*_*m*_, exhalation mask efficiency *M*_*ex*_, and quanta emission rate by one infected individual ER_*q*_.

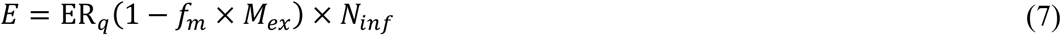

The first-order loss rate coefficient *λ* reflects several mechanisms: outdoor air ventilation *λ*_1_, filtration *λ*_2_, deposition on surfaces *λ*_3_, and airborne inactivation *λ*_4_.

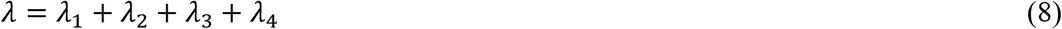

*λ*_1_ is the air change rate of outdoor air per hour (*h*^−1^) by the HVAC system or opening windows. *λ*_2_ is the in-room air filtration using portable air purifiers and/or duct filters in HVAC systems. *λ*_3_ is the removal by gravitational settling. Finally, *λ*_4_ is the inactivation/decay rate.

As already mentioned, the *PI*_*cond*_ is the conditional probability of infection, assuming that there is one infected person in the room. The prevalence rate of the disease in the city *P*_*prev*_ can be used to estimate the number of possible infected individuals in the room. Therefore, the absolute probability of infection *PI*_*abs*_ is calculated using the *PI*_*cond*_, *P*_*prev*_, and number of susceptible people in the room *N*_*sus*_ ^33^:

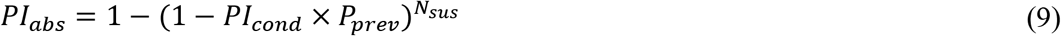

*N*_*sus*_ is calculated using the total number of people in the room *N*_*tot*_, number of infected persons, and the fraction of immune people in the community *F*_*immune*_. It can be estimated using the total recovered cases in the study region ^36,37^.

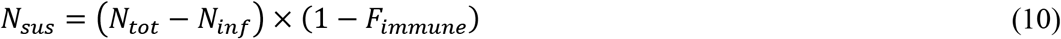

The prevalence rate of the disease is estimated using the daily COVID-19 statistics reported by official sources:

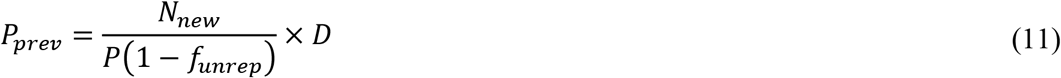

where *N*_*new*_ is the daily new cases, *P* is the population, *f*_*unrep*_ is the fraction of unreported cases. A study on ten diverse geographical sites in the US shows that the estimated number of infections was much greater than the number of reported cases in all sites ^36^. By doing more daily tests, the current unreported fraction is around 0.8. *D* is the duration of the infectious period of SARS-CoV-2 of 10 days in this work ^38^.

### 2.3 Sensitivity analysis model

Both CO_2_ and infection risk models include uncertain parameters such as ventilation rates and emission rates. Some of the uncertain parameters may impact the result’s accuracy and should be calibrated by measurement data. Ideally, with sufficient measurements and computer resources, all the uncertain parameters should be included in the calibration parameters. In reality, limited by data quality/quantity or computer resources, only a few parameters could be included. Many parameters and inputs could also manifest different levels of uncertainties and significances on simulation outputs. So it is impracticable and unnecessary to calibrate all parameters, but for dominant parameters only. Identifying these dominant parameters cannot merely rely on arbitrary parameter selections from modelers’ knowledge but should be based on a scientific process, i.e., a sensitivity analysis.

To conduct a sensitivity analysis process, prior distributions and ranges of selected unknown parameters should be determined. Then Monte Carlo (MC) simulation is employed to propagate simulations whose model parameters’ values are randomly chosen from the predefined ranges using a specific sampling method to perform simulation runs iteratively. Here, the Latin Hypercube Sampling (LHS) method ^39^ is applied since it provides good convergence of parameter space with relatively fewer samples. The obtained input-output dataset is then employed to identify the dominant model parameters that strongly affect the outputs. In this study, 440 parametric simulations were conducted for the sensitivity analysis of the CO_2_ concentration model ^40^.

The importance ranking results may vary with different combinations of sensitivity methods and outputs depending on the variety of fundamental algorithms and conditions of each sensitivity analysis method ^41^. To avoid the potential inconsistency, Lim and Zhai proposed a new sensitivity analysis method, sensitivity value index (SVI), to account for the differences in sensitivity analysis methods and target outputs ^42^. Eq. 12 shows how SVI is applied to recognizing and comparing the importance rankings from different sensitivity analysis methods through the normalization and aggregation process.

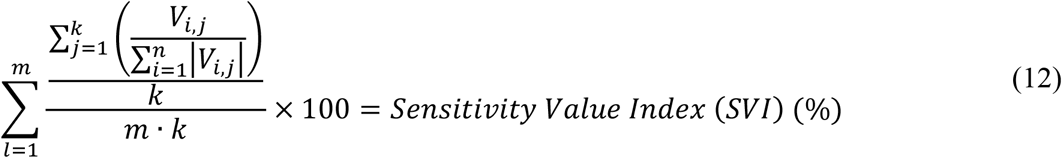

where *V*_*i,j*_ is the value of a sensitivity analysis method, *i* is a parameter, *n* is the total number of the parameters, *j* is a sensitivity method, *k* is the total number of sensitivity methods, *l* is the target output, and *m* is the total number of target outputs.

### 2.4 Bayesian calibration and Markov Chain Monte Carlo (MCMC) models

As the footstone of all Bayesian statistics, Bayes’ theorem was first proposed by Reverend Thomas Bayes in his doctoral dissertation ^43^ and can be described as:

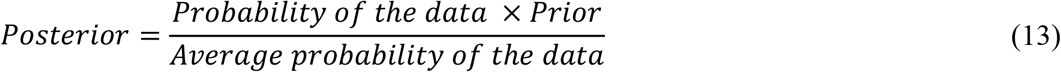

The probability of an event is inferred based on the prior knowledge of conditions related to the event. Bayesian inference is one application of Bayes’ theorem and can be written as:

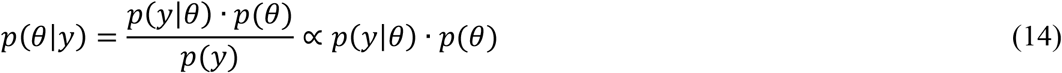

where *p*(*θ*|*y*) is the posterior distribution of the unknown parameter *θ* based on known observation *y. p*(*y*|*θ*) is the likelihood function of observation conditional on the unknown parameter. *p*(*θ*) is the prior distribution of the unknown parameter which is the marginal probability that means it is irrespective of the outcome of another variable, and *p*(*y*) is the probability of the observation that is marginal as well to normalize *p*(*y*|*θ*). Therefore, the posterior probability is proportional to the product of the prior probability and the likelihood.

In reality, it is impractical to apply the Bayesian inference for analytical solutions to all problems because the likelihood’s integrals can be computationally expensive or sometimes impossible to be calculated. MCMC is a versatile approach to solve the parameter estimation problem with two components. One is the well-known Monte Carlo method. It is a computational algorithm to solve statistically challenging problems relying on repeated random samplings and approximate the target value (e.g., mean value) using the independent samples’ results. The other is the Markov Chain method for solving a sequence of possible events. The probability of each event depends only on the state attained in the previous event. By combining MCMC and Bayesian inference, posterior distribution can be estimated efficiently.

Different MCMC algorithms can be classified into either a “random walking” group or a gradient-based group according to the acceptance-rejection criterion’s adoption. In this study, Hamiltonian Monte Carlo (HMC) sampling method ^44^ was used for the MCMC. Five thousand steps of the HMC algorithms on each of four separate chains were explored in this study to make a total of 20,000 samplers. We used one thousand samples during the “warming-up” stage to move chains toward the highest density area and tune sampler hyperparameters. For each room, the first 2/3 of measurements are used for the calibration, with the remaining for the validation.

In this study, the Coefficient of Variance of Root Mean Squared Error (CVRMSE) (Eq. 15) and Normalized Mean Bais Error (NMBE) were used as indicators to estimate the calibration and validation performance.

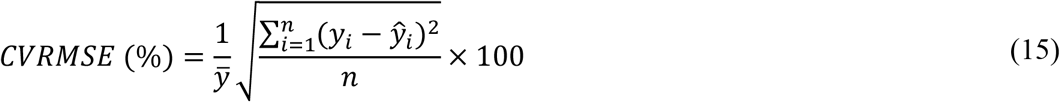

where *ŷ*_*i*_ is a predicted variable value for period *i, y*_*i*_ is an observed value for period *i*, 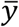 is the mean of the observed value, and *n* is the sample size.

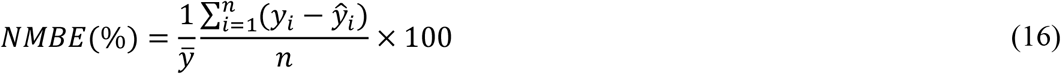

## 3. Case study

In this study, three typical classrooms of three different schools in Montreal, Canada, were selected for calibration and infection risk analysis. Each classroom was monitored during a typical pandemic day, and occupants’ information (students’ age and number), ventilation system status, window status, and transient indoor CO_2_ concentration are recorded. The summary information is shown in Table 1 and Figure 2.

**Table 1.** Basic information about the measured classrooms

| Room NO. | Date | Volume (m<sup>3</sup>) | MV | NV | Occupancy Number (student + adult) | Student age |
| --- | --- | --- | --- | --- | --- | --- |
| 1 | Nov. 10, 2020 | 165 | Y | Y | 19 + 1 | 7 |
| 2 | Nov. 6, 2020 | 236 | N | Y | 20 + 1 | 11 |
| 3 | Nov. 9, 2020 | 236 | N | Y | 18 + 1 | 7 |

Notes: MV means mechanical ventilation; NV means natural ventilation.

**Fig. 2.**
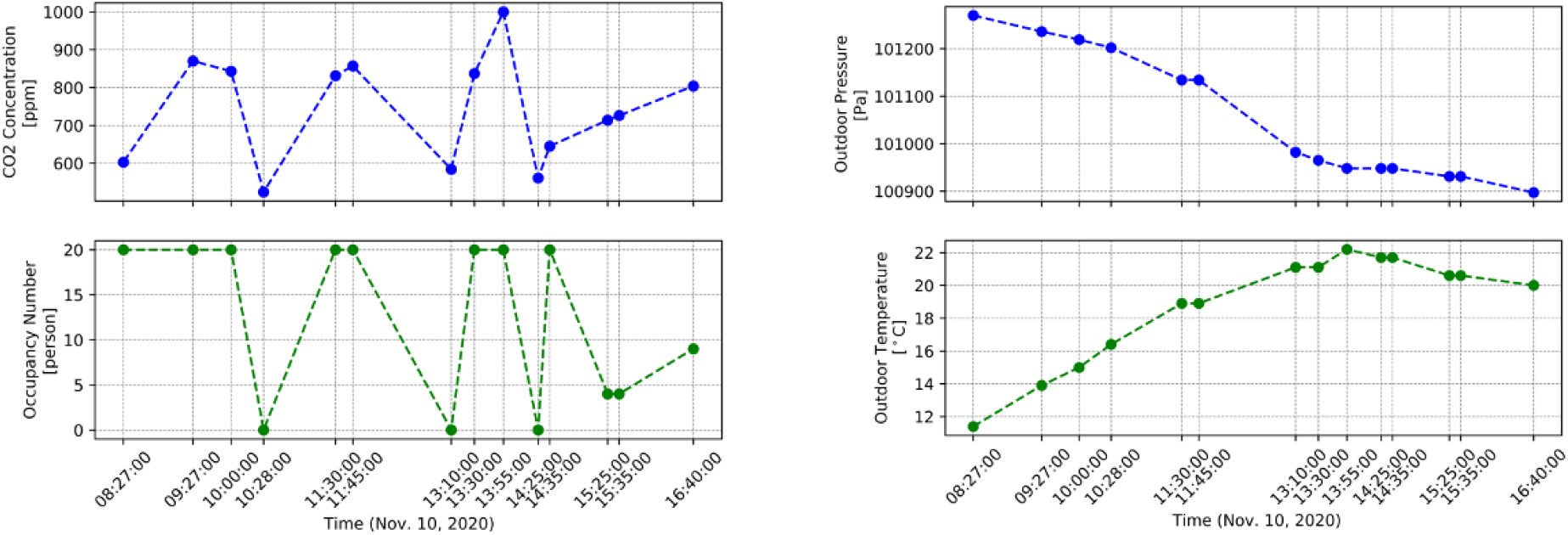
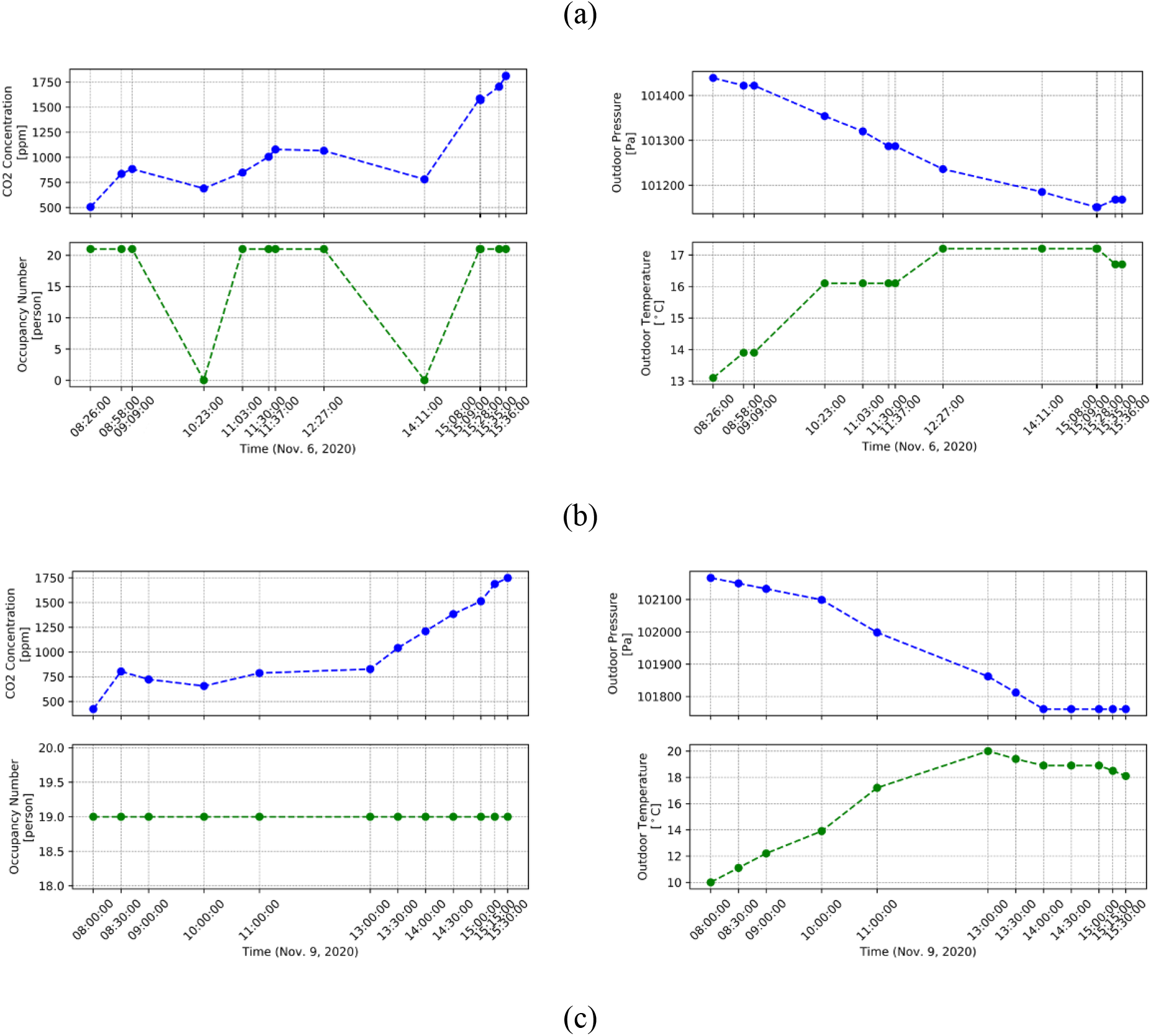
CO_2_ concentration measurements, occupancy number profile, outdoor air temperature, and outdoor air pressure of a) Room 1, b) Room 2, c) Room 3.

Room 1 is equipped with a mechanical ventilation system, and CO_2_ is between 500 ∼ 1000 ppm, while for Room 2 and Room 3, with no access to a mechanical ventilation system, the CO_2_ reaches up to 1800 ppm. The CO_2_ in Room 1 seems to indicate an acceptable level of air quality (<1000 ppm), and for Rooms 2 and 3, it is higher than the acceptable level of air quality requirements. The occupancy pattern of Room 1 and Room 2 was measured, but for Room 3, no occupancy pattern was recorded, so we assumed that number of students is constant as recorded in the morning during the day. The outdoor air temperature and pressure data are provided by Environment and Climate Change Canada ^45^. Other parameters required for the CO_2_ calculation, such as outdoor air CO_2_, generation rate, and outdoor air ventilation rate, are not available; therefore, we calibrate them using the CO_2_ measurements.

## 4. Results

In this section, the sensitivity analysis finds the dominant parameters for the calculation of CO_2_ concentrations. Then we use the Bayesian MCMC calibration to estimate the daily average ventilation rate using the CO_2_ measurement data and occupancy patterns. With the calibrated model, we calculate the CO_2_ concentrations in the three rooms under different ventilation rates. Finally, we used the calibrated ventilation rate to estimate the infection risk in each classroom. We also find the ventilation rate and CO_2_ level thresholds to avoid the airborne COVID-19 aerosol spread for different exposure times.

### 4.1 CO_2_ model sensitivity analysis

Outdoor/indoor pressure, outdoor/indoor air temperature, occupancy number, room volume, outdoor air ventilation rate, and CO_2_ exhale rate are input parameters to predict CO_2_ concentration. For the sensitivity analysis, the range of selected model inputs/parameters were defined according to the references, codes, and standards available. Table 2 shows the parameters with their sensitivity importance rankings: a smaller number indicates a more important/sensitive parameter.

**Table 2.**
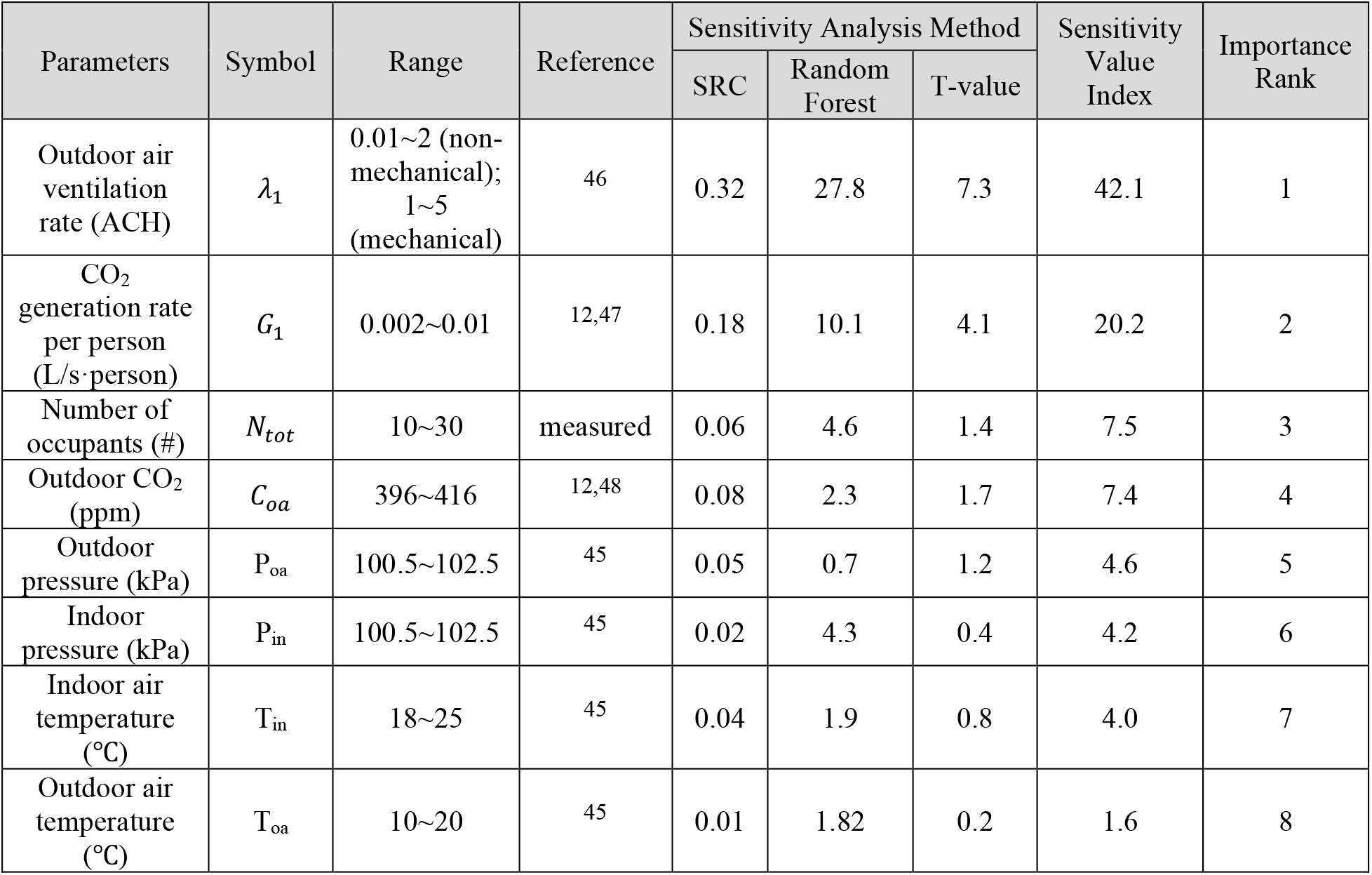
Sensitivity analysis with importance ranking for indoor CO_2_ concentration.

| Parameters | Symbol | Range | Reference | Sensitivity Analysis Method |  |  | Sensitivity Value Index | Importance Rank |
| --- | --- | --- | --- | --- | --- | --- | --- | --- |
|  |  |  |  | SRC | Random Forest | T-value |  |  |
| Outdoor air ventilation rate (ACH) | $\lambda_1$ | 0.01~2 (non-mechanical);<br>1~5 (mechanical) | 46 | 0.32 | 27.8 | 7.3 | 42.1 | 1 |
| CO<sub>2</sub> generation rate per person (L/s·person) | $G_1$ | 0.002~0.01 | 12,47 | 0.18 | 10.1 | 4.1 | 20.2 | 2 |
| Number of occupants (#) | $N_{tot}$ | 10~30 | measured | 0.06 | 4.6 | 1.4 | 7.5 | 3 |
| Outdoor CO<sub>2</sub> (ppm) | $C_{oa}$ | 396~416 | 12,48 | 0.08 | 2.3 | 1.7 | 7.4 | 4 |
| Outdoor pressure (kPa) | $P_{oa}$ | 100.5~102.5 | 45 | 0.05 | 0.7 | 1.2 | 4.6 | 5 |
| Indoor pressure (kPa) | $P_{in}$ | 100.5~102.5 | 45 | 0.02 | 4.3 | 0.4 | 4.2 | 6 |
| Indoor air temperature (°C) | $T_{in}$ | 18~25 | 45 | 0.04 | 1.9 | 0.8 | 4.0 | 7 |
| Outdoor air temperature (°C) | $T_{oa}$ | 10~20 | 45 | 0.01 | 1.82 | 0.2 | 1.6 | 8 |

It is concluded that, for the classroom CO_2_ levels, the most dominant parameters are outdoor air ventilation rate, CO_2_ generation rate per person, number of occupants, and outdoor CO_2_ concentration. Specifically, the outdoor air ventilation rate’s SVI is two times the second important parameter. Some sensitive parameters are often known, such as occupant number, outdoor temperature, and pressure. So they may not need to be calibrated. Therefore, we selected the outdoor air ventilation rate, CO_2_ generation rate, outdoor CO_2_ concentration, and indoor air temperature for the next step’s model calibration. Indoor pressure was assumed to be identical to the outdoor pressure.

### 4.3 Calibration and validation performance

For the calibration of the CO_2_ model, the Bayesian inference method was applied. For each occupancy phase (e.g., between every two measurements), we use the new measured CO_2_ data as the initial condition for Eq. 2. The posterior distributions are shown in Figure 3 and Table 3. In each subplot, the red dash line represents the parameter’s prior distribution in Table 2.

**Table 3.** Calibrated parameters of the CO_2_ model

| Room NO. | Prior Distribution | Posterior Distribution |  |  |  |  |  |  |
| --- | --- | --- | --- | --- | --- | --- | --- | --- |
|  | Uniform Distribution Range | Mean Value | Standard Deviation | Quantiles (%) |  |  |  |  |
|  |  |  |  | 2.5 | 25 | 50 | 75 | 97.5 |
| Outdoor Air Ventilation Rate (ACH) |  |  |  |  |  |  |  |  |
| Room 1 | (1, 5) | 1.96 | 0.16 | 1.67 | 1.85 | 1.96 | 2.07 | 2.29 |
| Room 2 | (0.01, 2) | 0.40 | 0.04 | 0.33 | 0.38 | 0.40 | 0.43 | 0.48 |
| Room 3 |  | 0.79 | 0.03 | 0.73 | 0.77 | 0.79 | 0.81 | 0.86 |
| CO<sub>2</sub> Generation Rate [$\times 10^{-3}$ L/s/person] | | | | | | | | |
| Room 1 | (0.002, 0.01) | 2.42 | 0.16 | 2.12 | 2.30 | 2.41 | 2.52 | 2.75 |
| Room 2 |  | 2.14 | 0.08 | 2.01 | 2.07 | 2.13 | 2.19 | 2.32 |
| Room 3 |  | 2.02 | 0.02 | 2.00 | 2.01 | 2.01 | 2.03 | 2.07 |
| Outdoor CO<sub>2</sub> Concentration (ppm) |  |  |  |  |  |  |  |  |
| Room 1 | (396, 416) | 414.4 | 1.62 | 410.1 | 413.8 | 414.9 | 415.6 | 416.0 |
| Room 2 |  | 406.5 | 5.72 | 396.6 | 401.7 | 406.9 | 411.6 | 415.5 |
| Room 3 |  | 401.8 | 4.89 | 396.2 | 397.9 | 400.4 | 404.7 | 414.1 |
| Indoor Air Temperature (K) |  |  |  |  |  |  |  |  |
| Room 1 | (18, 25) | 297.0 | 1.13 | 293.8 | 296.6 | 297.4 | 297.9 | 298.1 |
| Room 2 |  | 294.7 | 2.03 | 291.4 | 293.0 | 294.8 | 296.6 | 298.0 |
| Room 3 |  | 293.9 | 1.92 | 291.2 | 292.2 | 293.6 | 295.4 | 297.7 |

**Fig. 3.**
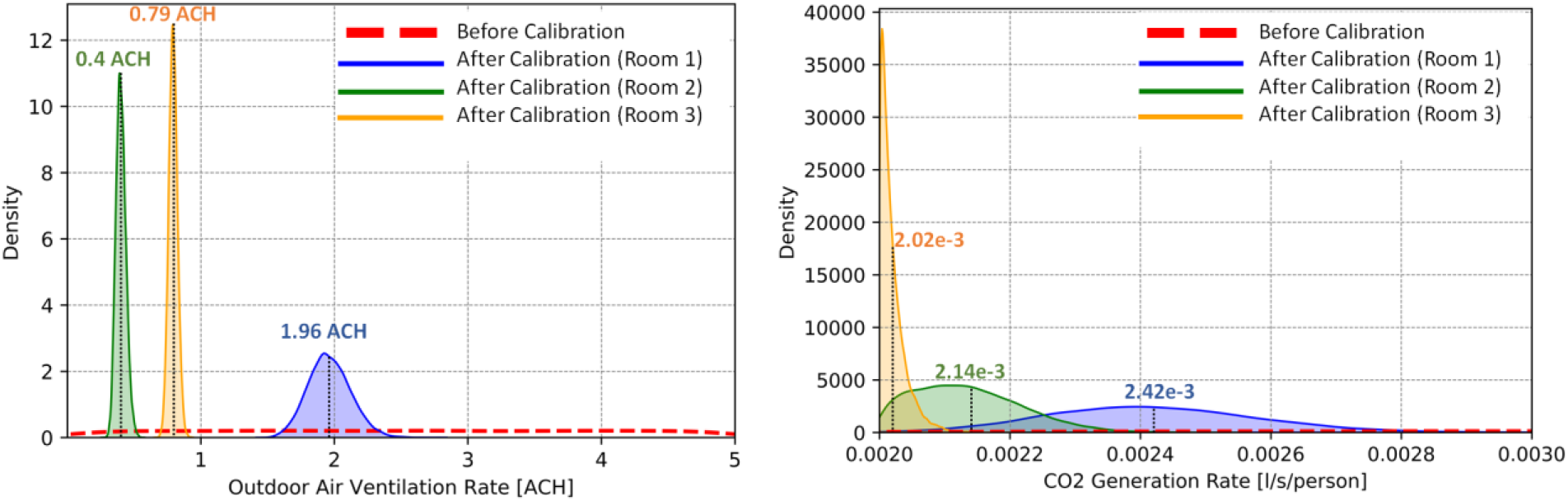
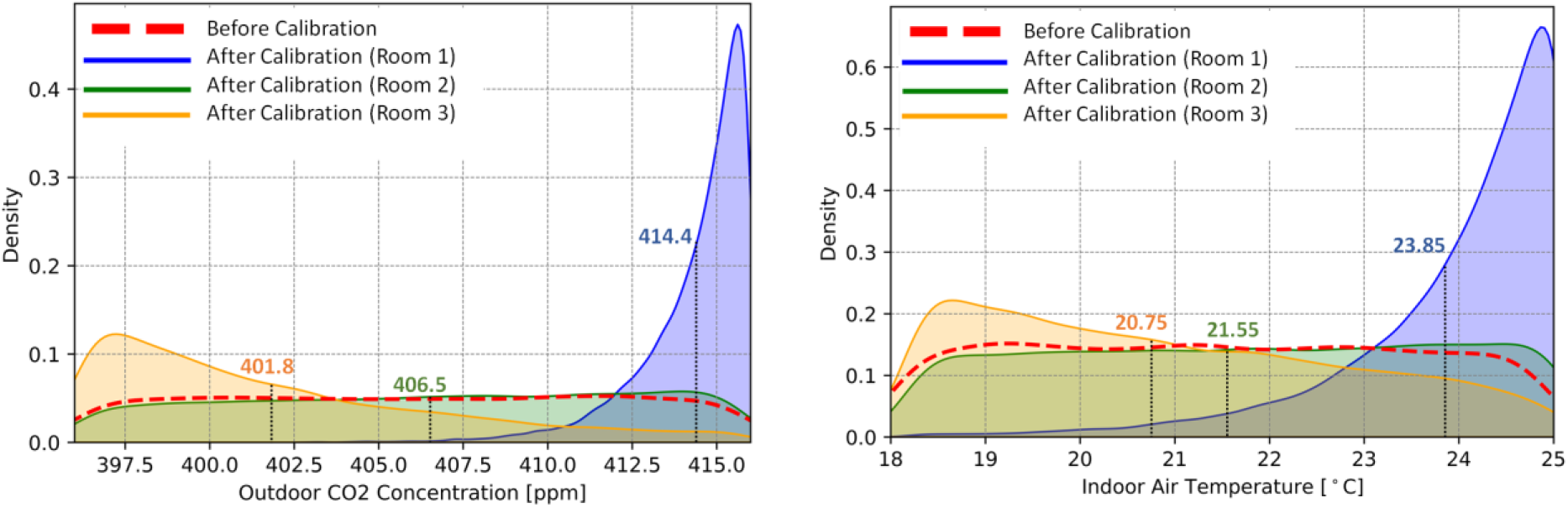
Distribution of calibrated parameters of the indoor CO_2_ model.

The posterior distributions with the Bayesian calibration are indicated by blue, green, and orange for Rooms 1, 2, and 3, respectively. The calibrated outdoor ventilation rate is 1.96 ± 0.31 ACH for Room 1, 0.40 ± 0.08 ACH for Room 2, and 0.79 ± 0.06 ACH for Room 3. Here, the ventilation rate is expressed by the calibrated mean value followed by the uncertainty for a 95% confidence interval. Room 1 is mechanically and naturally ventilated (i.e., open windows), so its ventilation rate is significantly higher than Rooms 2 and 3, with the outdoor air only from open windows. The results of Room 2 and Room 3 are closer since both are naturally ventilated with the same room volumes. The span of the posterior distribution of Room 1 is larger because of its wider prior distribution. For Room 3, a constant occupancy was used, so it is relatively easier for the MCMC iteration to find a posterior distribution. For the outdoor CO_2_ level and indoor air temperature, Room 2 and Room 3 are closer than Room 1 due to different ventilation modes.

Using the mean value of the calibration parameters, we compared the simulation results and measurements of CO_2_ in Figure 4. The simulation results show similar trends well as the measurements. According to the American Society of Heating, Refrigerating, and Air-conditioning Engineers (ASHRAE) guideline 14 ^49^ and FEMP ^50^, when the CVRMSE and NMBE values are less than 30% and ±10% for transient data, the calibrated accuracy meet the requirements. The (CVRMSE, NMBE) of validation of Rooms 1-3 are (15.3, 7.6), (10.5, 6.1), and (12.5, 12.3), respectively, which shows that the validated accuracy is satisfied.

**Fig. 4.**
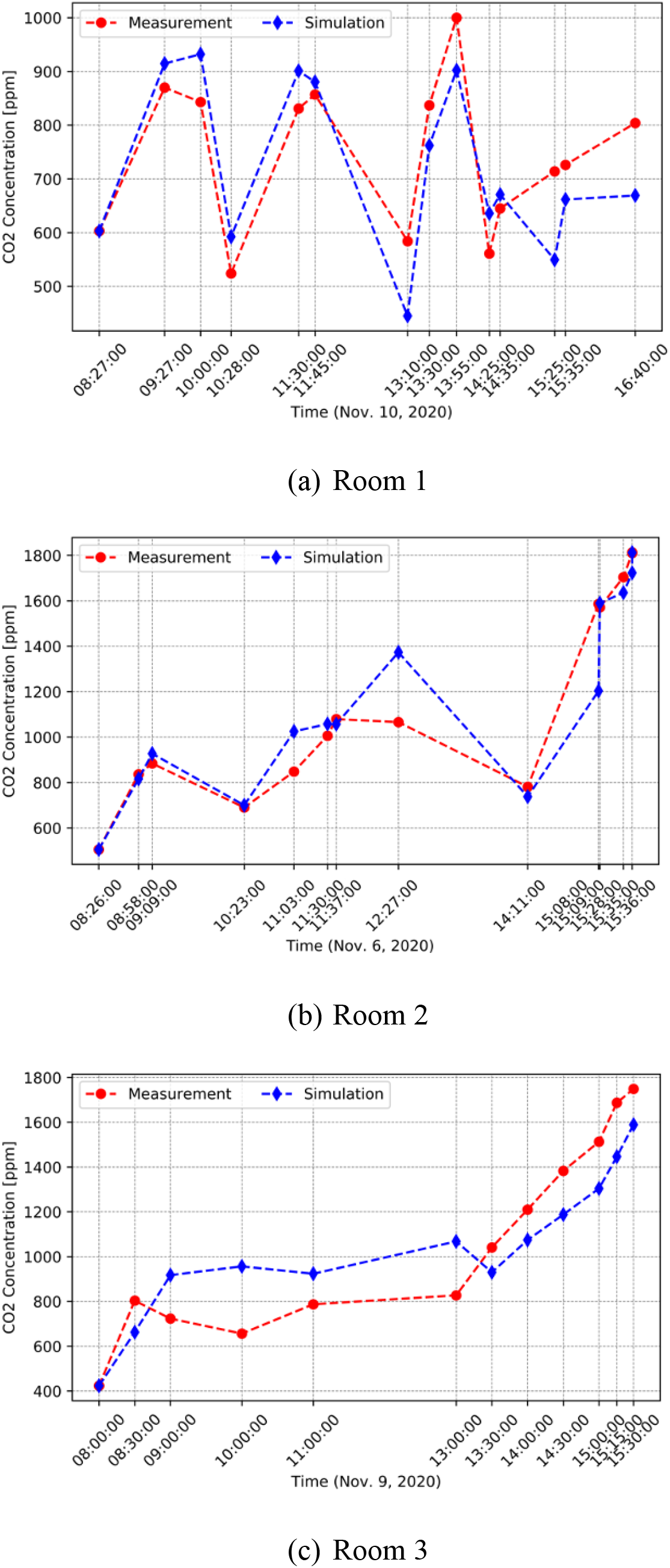
Comparison of simulated and measured CO_2_ levels in three schools.

### 4.4 CO_2_ level and ventilation rate evaluations

The calibrated ventilation rates in all three classrooms are all less than 2 ACH. The recommended ventilation rate for a safe indoor environment by Harvard-CU Boulder Portable Air Cleaner Calculator for Schools is at least 5 ACH. Therefore, the ventilation rate of all three classrooms seems inadequate. To relate the CO_2_ levels, e.g., measured from a CO_2_ sensor, and the ventilation rates, by using the calibrated CO_2_ model, we calculate the outdoor air ventilation rate in ACH and CFM/person as a function of CO_2_ levels at different exposure times in Figure 5. It helps teachers find the room ventilation rate directly based on the CO2 sensors at different school hours. For example, for Room 1, when CO_2_ > 600 ppm, OA (outdoor air) < 5 ACH (24 CFM/person); CO_2_ > 800 ppm, OA < 2 ACH (9 CFM/person) at any time of the day. A CO_2_ level less than 480 ppm indicates a ventilation rate greater than 10 ACH (48 CFM/person) at all times. For Room 2, the same CO_2_ levels correspond to a lower ventilation rate than Room 1. For example, when CO_2_ > 600 ppm, OA < 2 ACH (13 CFM/person); CO_2_ < 440 ppm indicates that OA > 10 ACH (66 CFM/person). This shows that Room 1 shows a higher ventilation rate (ACH) for the same CO_2_ level because of its smaller size. The ventilation rate of Room 3 at a specific CO_2_ level is higher than Room 2 because of its constant occupancy compared to the variable occupancy of Room 2. It seems lunch breaks indeed lower CO_2_ levels significantly (thus infectious risk in schools). For Room 3, CO_2_ > 600 ppm indicates OA < 3 ACH (22 CFM/person). These results show that the indoor CO2 could vary significantly in different classrooms even with the same ventilation rate because of different room sizes, occupants number, and occupancy schedule. All three classrooms show that an indoor CO_2_ lower than 450 ppm indicates a ventilation rate greater than 10 ACH, close to the recommended 12 ACH value to prevent airborne transmission in health-care facilities^7,8^. Note that here CO_2_ is the exposure-time-averaged instead of the instantaneous level.

**Fig. 5.**
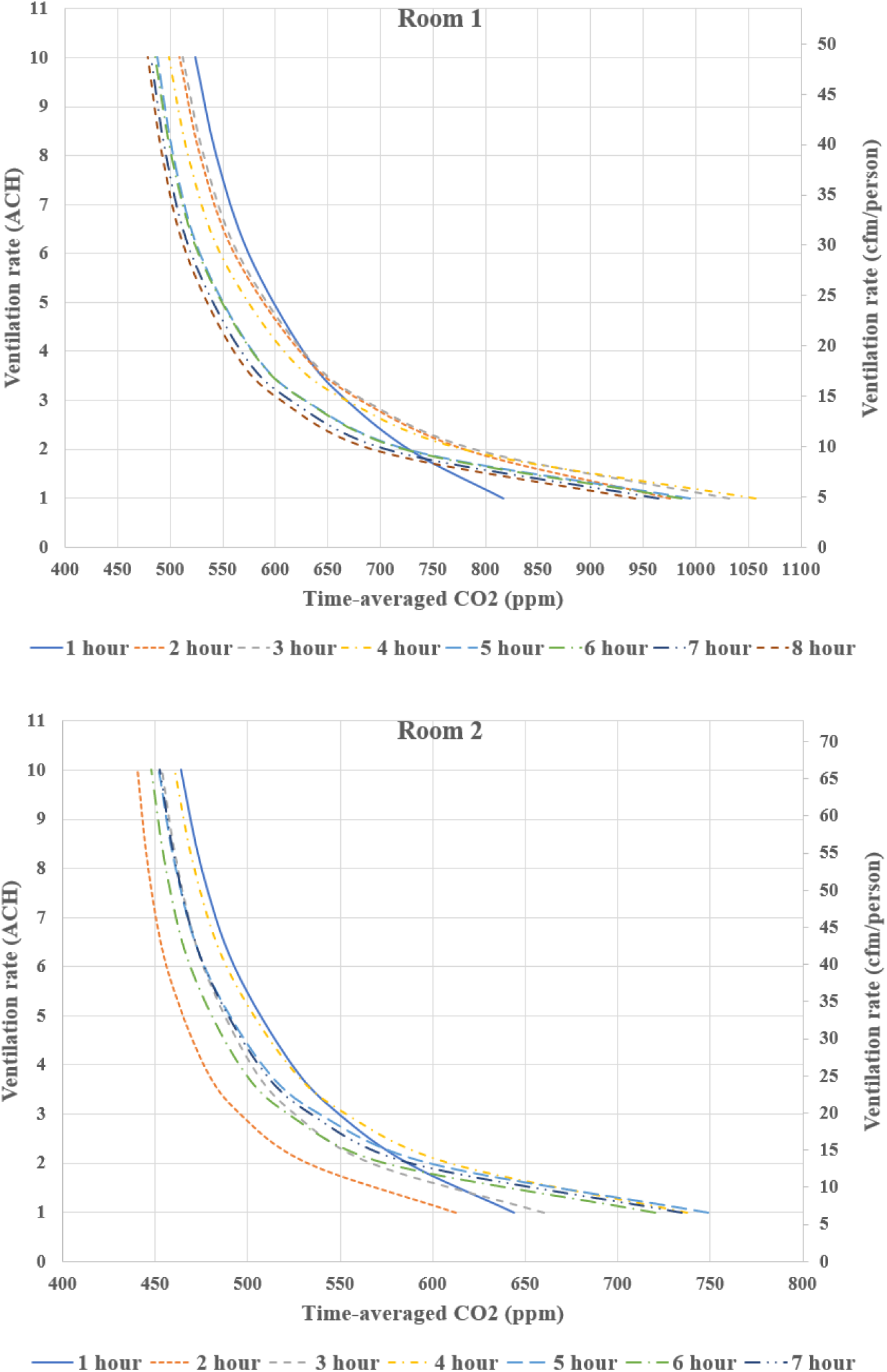
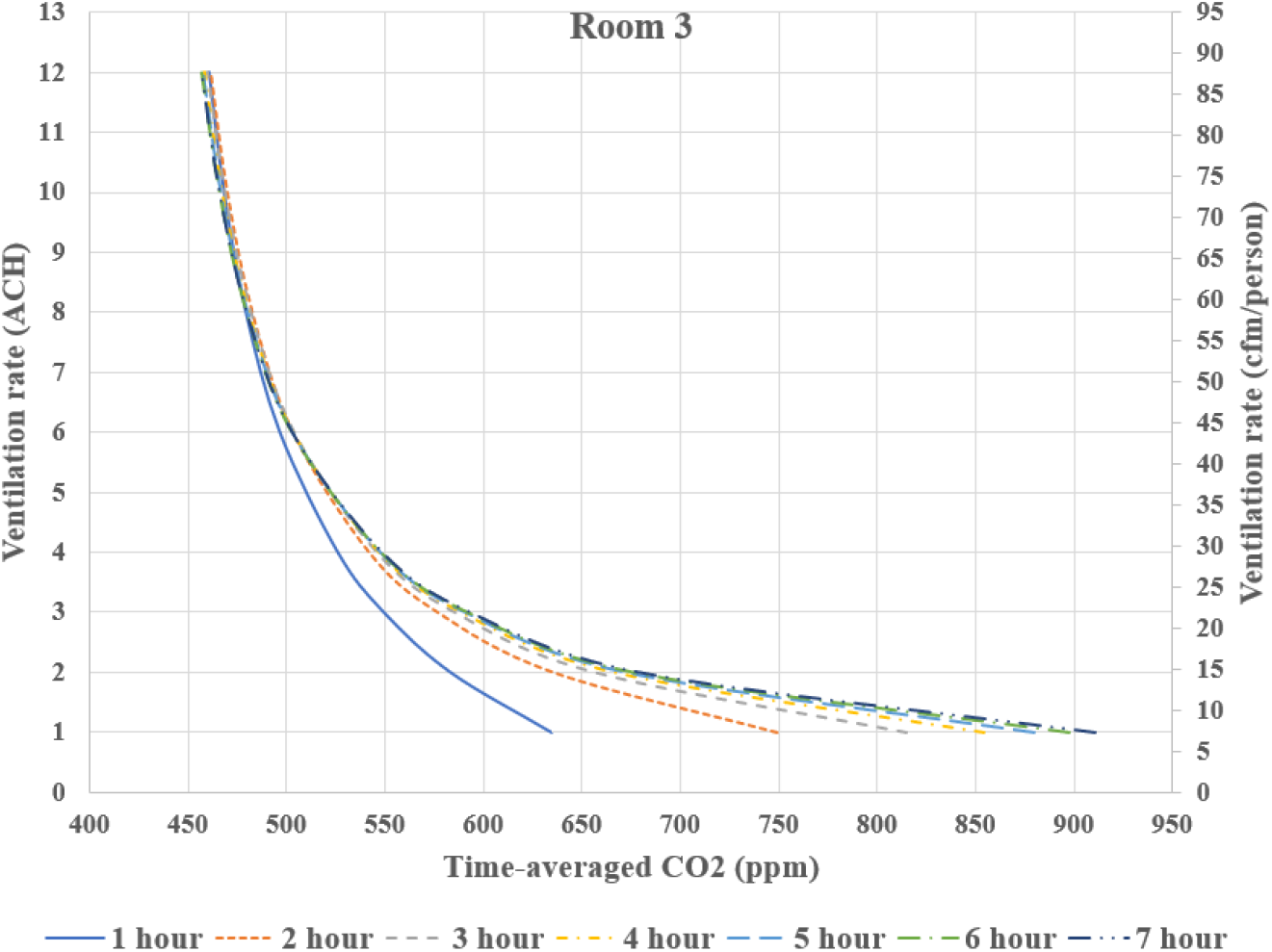
Relation of indoor CO_2_ levels and ventilation rates for classrooms.

### 4.5 CO_2_ level and infection risk evaluations

To study CO_2_ and airborne aerosol infectious risk relation, we first calculate the probability of infection risk with the posterior distribution of the ventilation rate obtained in Section 4.3. Note, Here, the Bayesian and MCMC analysis allow us to quantify the uncertainties of the ventilation rates to estimate airborne infectious risk by defining the probability of the infection risk: the probable range of the infection risk estimated in classrooms. Then, we evaluate different ventilation rates to identify the corresponding CO_2_ threshold level, at which the reproductive number, R_0_ < 1, at all exposure times. We estimate the infection risk and indoor air CO_2_ threshold based on the actual room conditions in this work. The recommended threshold could be used for other rooms under a similar condition.

Table 4 shows the input parameters for Eqs. 3-11 to calculate the COVID-19 airborne infection risk in classrooms. Actual room conditions with age and activity levels were used to determine breathing and quanta emission rates ^51,52^. All students wore a face mask in the classroom, and mask efficiency was selected based on students’ typical mask type. The prevalence rate and immunity fraction were calculated based on the official reported data^53^.

**Table 4.** Input parameters for calculation of infection risk in three classrooms

| Parameter | Symbol (unit) | Room number |  |  |
| --- | --- | --- | --- | --- |
|  |  | Room 1 | Room 2 | Room 3 |
| Breath rate | $B \text{ (m}^3/\text{h)}$ | 0.67 | 0.67 | 0.67 |
| Mask fraction | $f_m$ | 1 | 1 | 1 |
| Mask inhalation efficiency | $M_{in}$ | 30% | 30% | 30% |
| Mask exhalation efficiency | $M_{ex}$ | 50% | 50% | 50% |
| Quanta emission rate | $ER_q \text{ (q/h)}$ | 27.55 | 27.55 | 27.55 |
| Number of infected people | $N_{inf}$ | 1 | 1 | 1 |
| Indoor air filtration rate | $\lambda_2 \text{ (1/h)}$ | 0 | 0 | 0 |
| Deposition rate | $\lambda_3 \text{ (1/h)}$ | 0.3 | 0.3 | 0.3 |
| Decay rate | $\lambda_4 \text{ (1/h)}$ | 0.62 | 0.62 | 0.62 |
| Number of susceptible | $N_{sus}$ | 19 | 20 | 18 |
| Recovered cases | — | 38891 | 38014 | 38646 |
| Daily new cases | $N_{new}$ | 253 | 287 | 285 |
| Unreported cases | $f_{unrep}$ | 80% | 80% | 80% |
| Duration of disease | $D$ | 10 | 10 | 10 |
| Prevalence rate | $P_{prev}$ | 0.62% | 0.70% | 0.69% |

Figure 6 plots the transient PI_cond_ and PI_abs_ in three classrooms with the calibrated ventilation rates’ posterior distribution. The orange/green line is the baseline of PI_cond_/PI_abs_ calculated using the mean value of the posterior distribution of ventilation rate. The orange/green area is the estimated infection risk probability with a 95% confidence interval. For Rooms 1, 2, and 3, the mean *PI*_*cond*_ at the end of the day is around 14%, 14%, and 20%, respectively. Rooms 1 and 2 show lower *PI*_*cond*_ because no students were present during the break, so the quanta generation was zero. The uncertain band of *PI*_*cond*_ in Room 1 is wider than other rooms because of the larger posterior range of ventilation rate due to its mechanical ventilation system. In Rooms 1 and 2, when students leave the classroom, the *PI*_*abs*_ are zero because there is no susceptible persons in the room. Room 1 shows the lowest *PI*_*abs*_ because of the lower *PI*_*cond*_ and prevalence rate: all three measurements were conducted on different days. There are more students in Room 2 than Room 3; however, because of the lower *PI*_*cond*_, the absolute *PI* of Room 2 is thus lower.

**Fig. 6.**
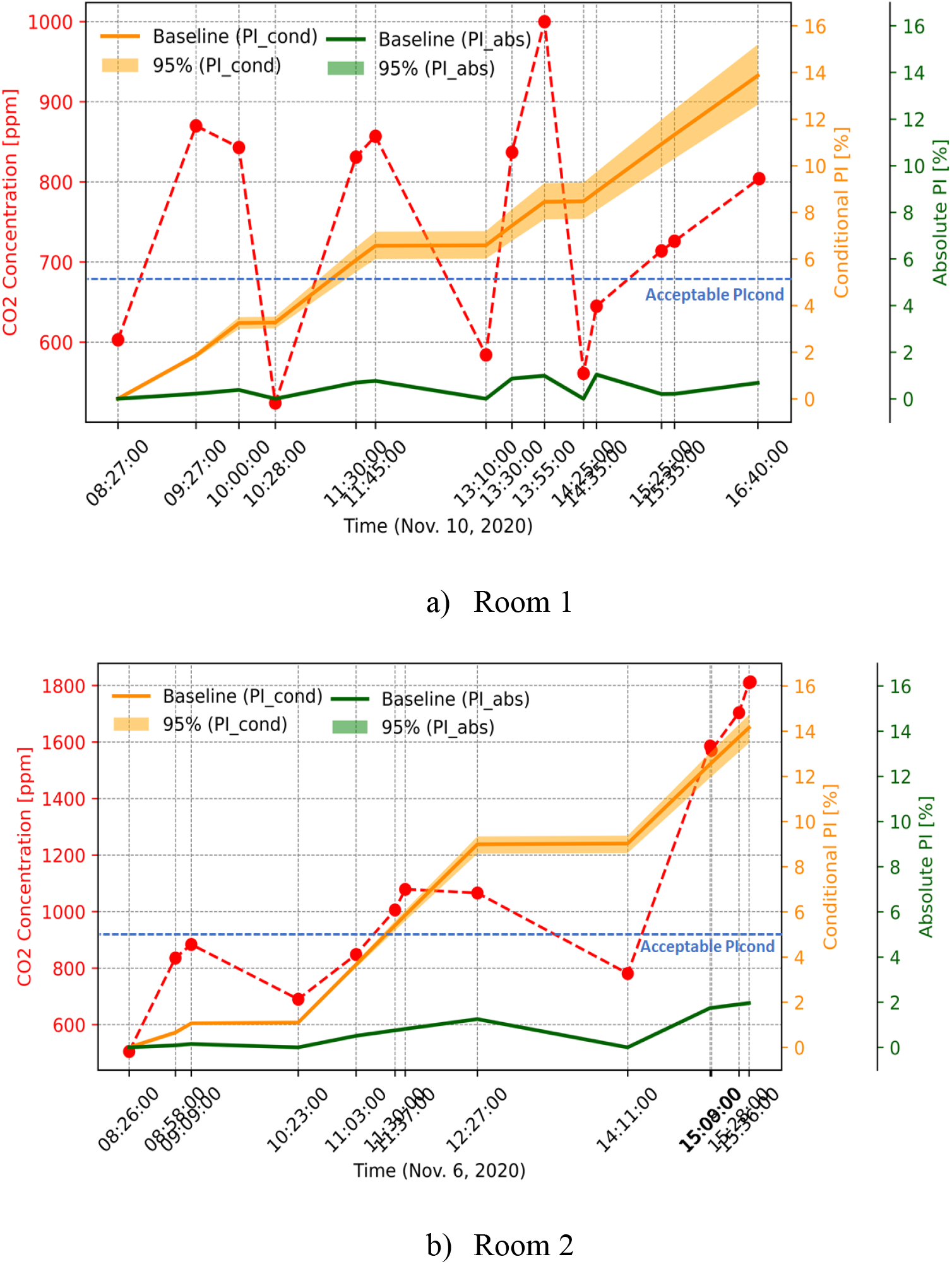
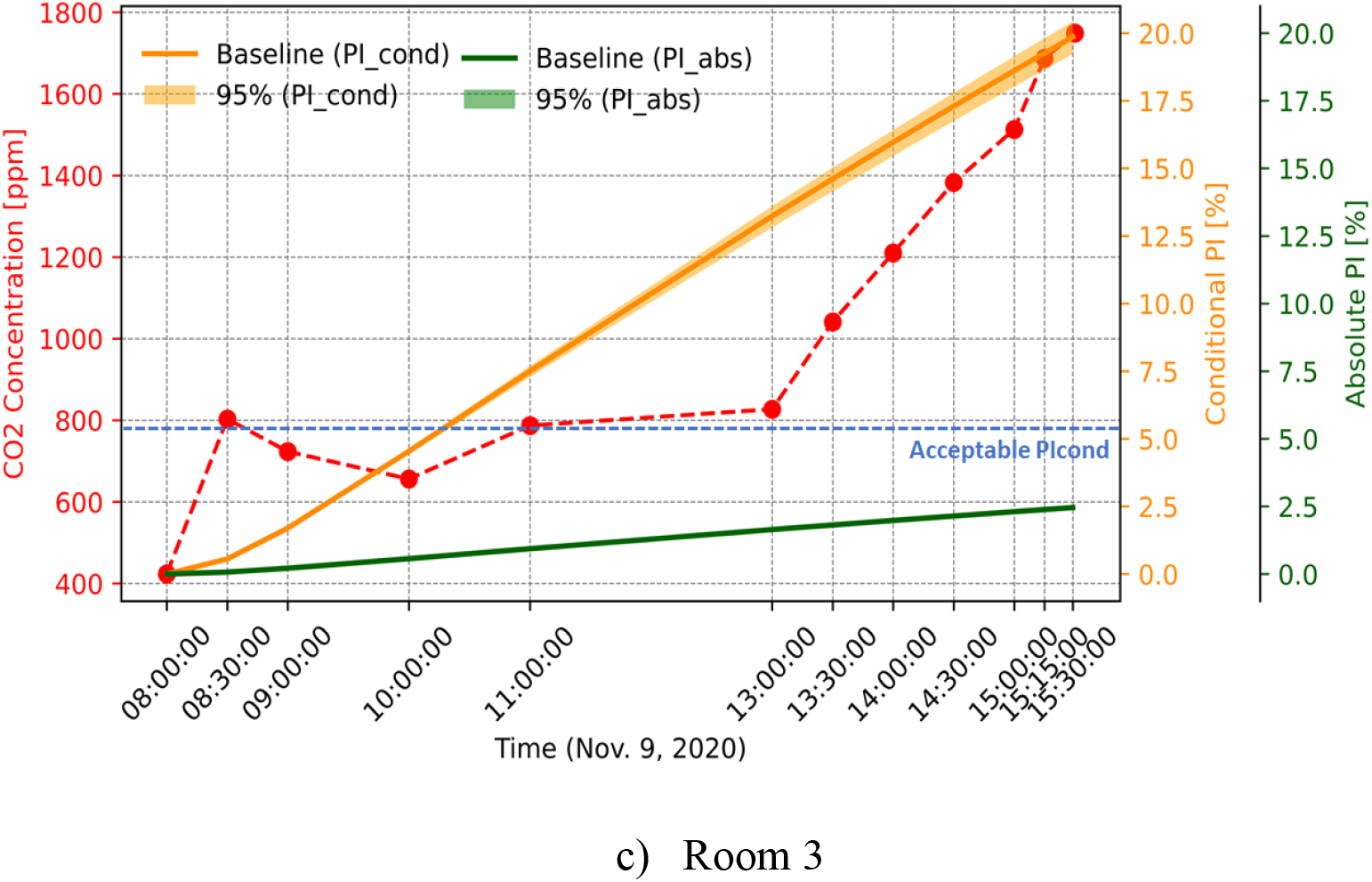
Probability of airborne infection risks in classrooms.

It also shows that after three hours, *PI*_*cond*_ of Room 1 is always smaller than two other rooms due to a higher ventilation rate. More frequent leaves of students (three breaks in a day) also contribute: the quanta level is generally lower than the two other rooms. *PI*_*cond*_ of Room 3 continues to rise with the exposure time, unlike other classrooms, again due to the constant occupancy assumed. Therefore, to reduce infection risk, more frequent breaks and leaves could be beneficial when increasing the classroom’s ventilation rate is relatively harder to achieve.

The conditional *PI* is the ratio of the number of infections to susceptibles *PI*_*cond*_ = *D*/*S*. The reproductive number (R_0_), defined by Rudnick and Milton (2003), is the number of secondary infections when a single infected person is introduced in the room, and everyone in the room is susceptible. If R_0_ < 1, then the infectious agent cannot spread in the population. For these three classrooms if *PI*_*cond*_ is smaller than 5.3%, 5%, and 5.5%, it is expected that the community spread in the classrooms could be stopped. Figure 6 shows that, for Rooms 1, 2, and 3, the conditional PI exceeds the level of R_0_ = 1 at around two ∼ three hours, after which mitigation measures should be taken.

To identify the relation between the required ventilation rates, corresponding CO_2_ levels, and the COVID-19 airborne spreading risks, we calculated them for the three classrooms as shown in Figure 7. At all exposure times, the indoor CO_2_ and *PI* decrease with an increased ventilation rate. The ventilation rate threshold to prevent the spread (R_0_<1) at all exposure times is 8, 3, and 6 ACH for Rooms 1, 2, and 3, respectively. Room 1 requires a higher ventilation rate because of the smaller room size. Room 2 needs a lower ventilation rate than Room 3 because students left the classroom several times. Therefore, the ventilation rate threshold depends on the occupancy schedule and the size of the room.

**Fig. 7.**
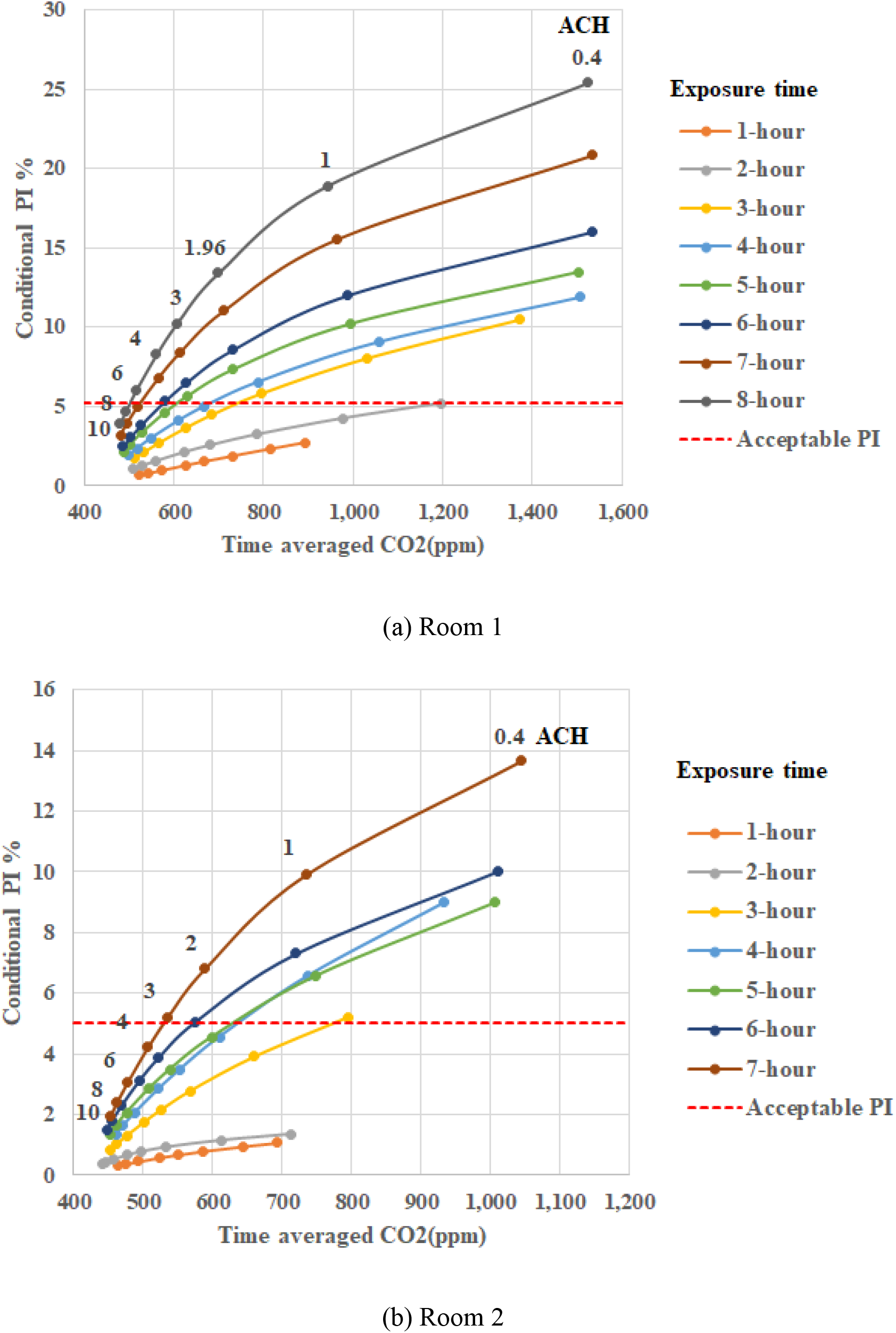
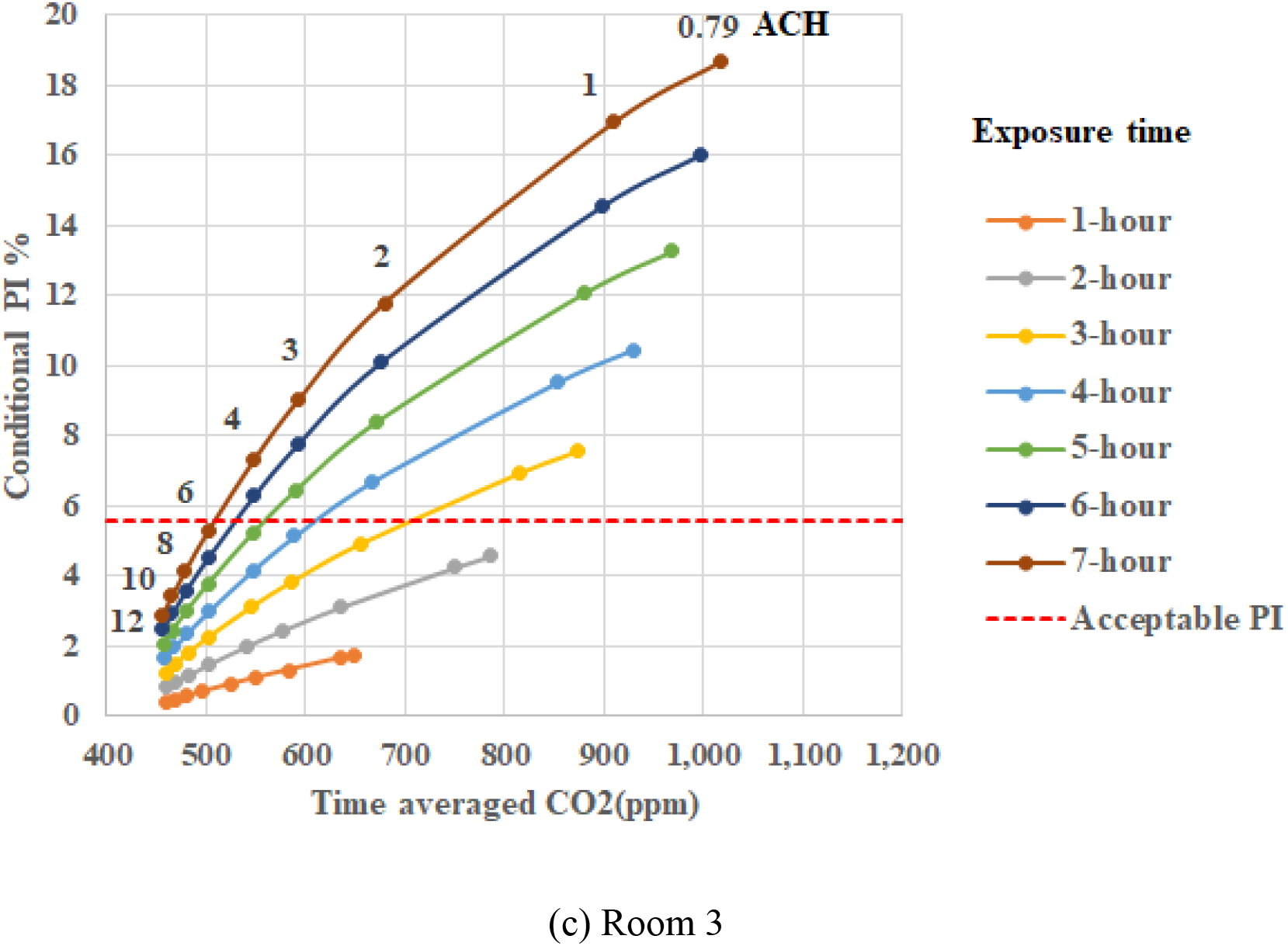
*PI*_*cond*_ and time-averaged CO_2_ for different exposure times and ventilation rates.

According to Figure 7, each classroom needs a different ventilation rate at different exposure times to reach the level of R_0_ = 1. On the other hand, we can find the indoor CO_2_ threshold for each exposure time corresponding *R*_*0*_ = 1, as shown in Figure 8. The CO_2_ threshold level decreases with exposure time for all three rooms. For Room 1 at the second hour, the acceptable level is around 1000 ppm and later has to be 520 ppm at the end of the day to avoid spreading. For Rooms 2 and 3, it decreases from 660 ppm to 505 ppm and 750 ppm to 500 ppm, respectively. Meanwhile, the threshold levels for 3-7 hours exposure times are close for all three classrooms. In comparison, Figures 8b and 8c illustrate that the ventilation rate threshold (to prevent the spreading) increases with exposure time and is not a constant number because of the three rooms’ different sizes and schedules.

**Fig. 8.**
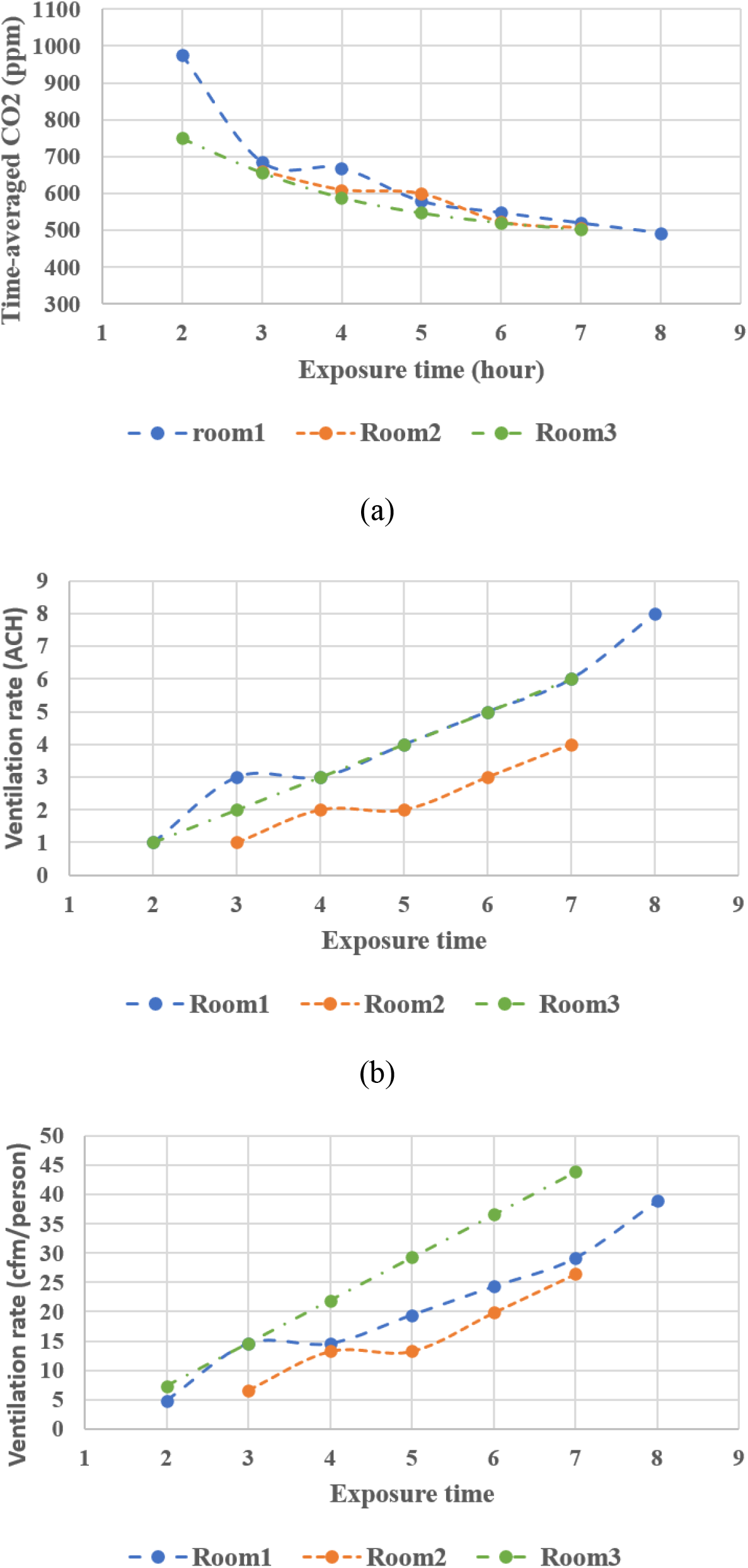
a) Time-averaged indoor CO_2_ concentration thresholds, and b, c) outdoor ventilation rate thresholds with exposure times to prevent spreading.

In summary, the results of all three classrooms show that the ventilation rate threshold to prevent the airborne transmission of COVID-19 depends on several parameters such as room size, student schedules, and exposure time. The indoor CO_2_ threshold seems to depend on exposure time mostly, and the time-averaged level of 500 ppm seems acceptable for all three classrooms.

## 5. Conclusion

The airborne transmission of COVID-19 is a major infection route in indoor spaces, especially with poor ventilation conditions, large occupancy density, and high exposure time, such as school classrooms. There are some recommended ventilation rates for acceptable indoor air quality or preventing airborne transmission in indoor spaces, but it is not easy to measure the actual room’s ventilation rate. Indoor air CO_2_ level can be used as an indicator for the ventilation rate, whereas it depends on several parameters that must be estimated. In this work, we used the sensitivity analysis, MCMC Bayesian Calibration method, measured data of indoor CO_2_ and occupancy profile of three classrooms in Montreal, Canada, to find and calibrate the dominant parameters to estimate CO_2_ levels indoors. The results showed that the outdoor ventilation rate is the most significant parameter. The calibrated ventilation rate with a 95% confidence level is 1.96 ± 0.31 ACH for Room 1 with mechanical ventilation, and 0.40 ± 0.08 ACH and 0.79 ± 0.06 ACH for Rooms 2 and 3 with windows open only. Based on the calibrated model, we created the correlations between the CO_2_ levels and the ventilation rates, which help teachers to estimate the ventilation rates from the CO2 sensors at any time. A time-averaged CO_2_ lower than 450 ppm is equivalent to a ventilation rate greater than 10 ACH in all three rooms, close to the recommended 12 ACH value for a safe indoor environment against airborne transmission.

This study also proposed the approach to calculating the probability of the infection risk based on the calibrated ventilation rate, which helps quantify the uncertainty of outdoor ventilation rates. Moreover, this study estimated the required ventilation rate threshold and the CO_2_ threshold values to prevent the airborne aerosol spreading of COVID-19 as a function of exposure time in the classrooms. The ventilation threshold at all hours is 8, 3, and 6 ACH for rooms 1, 2, and 3, respectively, and the CO_2_ threshold is around 500 ppm at all exposure times (< 8 hr) of a school day for all three classrooms. This threshold is significantly different from the recommended value of 1000 ppm for acceptable indoor air quality conditions. Therefore, it is recommended that the ventilation rate and indoor CO_2_ concentration thresholds be reconsidered in indoor spaces, especially classrooms, in the current pandemic.

## Data Availability

The data that support the findings of this study are available from the corresponding author, LW, upon reasonable request.

## Acknowledgment

The research is supported by the Natural Sciences and Engineering Research Council (NSERC) of Canada through the Discovery Grants Program [#RGPIN-2018-06734] and the Advancing Climate Change Science in Canada Program [#ACCPJ 535986-18].

